# Hospital length of stay for severe COVID-19 patients: implications for Remdesivir’s value

**DOI:** 10.1101/2020.08.10.20171637

**Authors:** Michaela R Anderson, Peter B. Bach, Matthew R. Baldwin

## Abstract

Remdesivir has been granted emergency use authorization for treatment of severe COVID-19. Remdesivir's pricing is based on a presumed reduction of hospital length of stay (LOS) by four days. But the Adaptive COVID-19 Treatment Trial (ACTT-1) that suggested this treatment benefit excluded patients who were expected to be discharged within 72 hours. Perhaps as a result, median time to recovery was unusually long in both arms of the study (15 days vs 11 days). Remdesivir requires a 5-day inpatient stay, so patients who would otherwise be discharged in fewer than 5 days may remain hospitalized to complete treatment while patients who would be discharged between 5 and 8 days, would only have potential reductions in their hospital LOS of 0-3 days. In a retrospective analysis of 1643 adults with severe COVID-19 admitted to Columbia University Medical Center and the Allen community hospital between March 9, 2020 and April 23, 2020, median hospital LOS was 7 (3-14) days. Five-hundred and eighty-six patients (36%) had a LOS of 1-4 days, 384 (23%) had a LOS of 5-8 days, and 673 (41%) were hospitalized for greater than or equal to 9 days. Remdesivir treatment may not provide the LOS reductions that the company relied on when pricing the therapy: 36% of the cohort would need to have LOS prolonged to receive a 5-day course, and only 41% of patients in our cohort had LOS of 9 days or more, meaning they could have their LOS shortened by 4 days and still receive a full Remdesivir course. Further investigation of shorter treatment courses and programs to facilitate outpatient intravenous Remdesivir administration are needed.

---

In June 2020 Gilead Sciences CEO announced that US hospitals would be charged $3,120 for a course of remdesivir, a price roughly 33% higher than the company plans to charge other high-income countries.^1^ The sole justification offered was that remdesivir would save hospitals $12,000 per patient by shortening hospital length of stay (LOS) by four days. This presumed benefit was extrapolated from the ACTT-1 trial, which found median time to recovery (an approximation of median time to hospital discharge) of 11 days in patients receiving remdesivir versus 15 days in those receiving placebo.^2^

But as justification for such a high price it is critical that the ACTT-1 finding is robust. It may not be, either because the outcomes of the study population are not generalizable, or because of exclusion from the trial of patients who were expected to be discharged within 72 hours. Remdesivir treatment requires a 5-day inpatient stay, so patients who would otherwise be discharged sooner may remain hospitalized to complete treatment. This also limits the achievable LOS reduction for patients who would otherwise be hospitalized 5-8 days. The remdesivir Emergency Use Authorization does not limit its use based on LOS, and specifies that intravenous remdesivir be given in an in-patient setting.^3^ We assessed what percentage of severe COVID-19 patients belonged to each of these LOS groups in a real world cohort.

## Methods

Our study cohort consisted of adults age ≥18 years with severe COVID-19 consecutively hospitalized from the emergency department (ED) at NewYork-Presbyterian Columbia University Medical Center and the Allen community hospital between March 9, 2020 and April 23, 2020, with follow-up through June 10, 2020. Data abstraction methods have been published previously.^4^ We defined severe COVID-19 based on remdesivir trial and FDA criteria: an initial oxygen saturation ≤94% on room air or the use of any supplemental oxygen within 24 hours of ED presentation.^2,5^ LOS was measured from presentation to death or hospital discharge.

## Results

There were 1,643 adults admitted with severe COVID-19, after excluding 21 (1%) who were discharged in <9 days to another hospital. The median age was 67 (interquartile range, 56-78) years, a majority were Hispanic or Black and had ≥1 comorbidity, 12% required mechanical ventilation within 24 hours, median LOS was 7 (3-14) days, and in-hospital 28-day mortality was 26%. Five-hundred and eighty-six patients (36%) had a LOS of 1-4 days, 384 (23%) had a LOS of 5-8 days, and 673 (41%) were hospitalized ≥9 days (Table). The distribution was similar when excluding patients who died during their hospital stay (Table). The majority of those with a LOS of 1-4 or 5-8 days were ≥60 years old (67% and 70%, respectively, Figure).

**Table:** Baseline characteristics by hospital length of stay

|  | Length of<br>stay 1-4<br>days<br>(n=586) | Length of<br>stay 5-8<br>days<br>(n=384) | Length of<br>stay ≥9<br>days<br>(n=673) | p value |
| --- | --- | --- | --- | --- |
| Median age, years (IQR) | 68 (55-80) | 68 (57-79) | 67 (56-75) | 0.03 |
| Sex, male | 325 (55) | 212 (55) | 424 (63) |  |
| Race/Ethnicity |  |  |  | 0.67 |
| White Non-Hispanic | 44 (8) | 33 (9) | 63 (9) |  |
| Black Non-Hispanic | 60 (10) | 41 (11) | 78 (12) |  |
| Hispanic | 318 (54) | 198 (52) | 329 (49) |  |
| Other | 83 (14) | 65 (17) | 104 (15) |  |
| Declined | 81 (14) | 47 (12) | 99 (15) |  |
| Comorbidities |  |  |  |  |
| Asthma/COPD | 84 (14) | 70 (18) | 125 (19) | 0.10 |
| Hypertension | 297 (51) | 194 (51) | 339 (50) | 0.99 |
| Chronic kidney disease | 79 (13) | 79 (21) | 123 (18) | 0.009 |
| Cancer | 53 (9) | 42 (11) | 102 (15) | 0.003 |
| Smoking | 42 (17) | 52 (14) | 83 (12) | 0.002 |
| Diabetes | 209 (36) | 153 (42) | 280 (42) | 0.045 |
| Pulmonary Heart Disease | 33 (6) | 24 (6) | 45 (7) | 0.74 |
| Median number of comorbidities (IQR) | 1 (0-2) | 1 (0-3) | 1 (0-3) | 0.005 |
| Participation in Remdesivir trial* | 7 (1) | 3 (1) | 35 (5) | <0.001 |
| Discharge Disposition |  |  |  | <0.001 |
| Home | 356 (61) | 238 (62) | 219 (33) |  |
| Rehabilitation Center | 35 (6) | 47 (12) | 190 (28) |  |
| Death | 168 (29) | 81 (21) | 173 (26) |  |
| Hospice | 27 (5) | 18 (5) | 30 (4) |  |
| Transferred to other hospital | 0 (0) | 0 (0) | 8 (1) |  |
| Hospitalized at study completion | 0 (0) | 0 (0) | 53 (8) |  |

**Definition of abbreviations:** COPD: chronic obstructive pulmonary disease; IQR: interquartile range. Values presented as n (%). *P* values calculated using Kruskal-Wallis tests or Chi-square tests.
\*Gilead GS-US-540-5774 or GS-US-540-5773.

**Figure:**
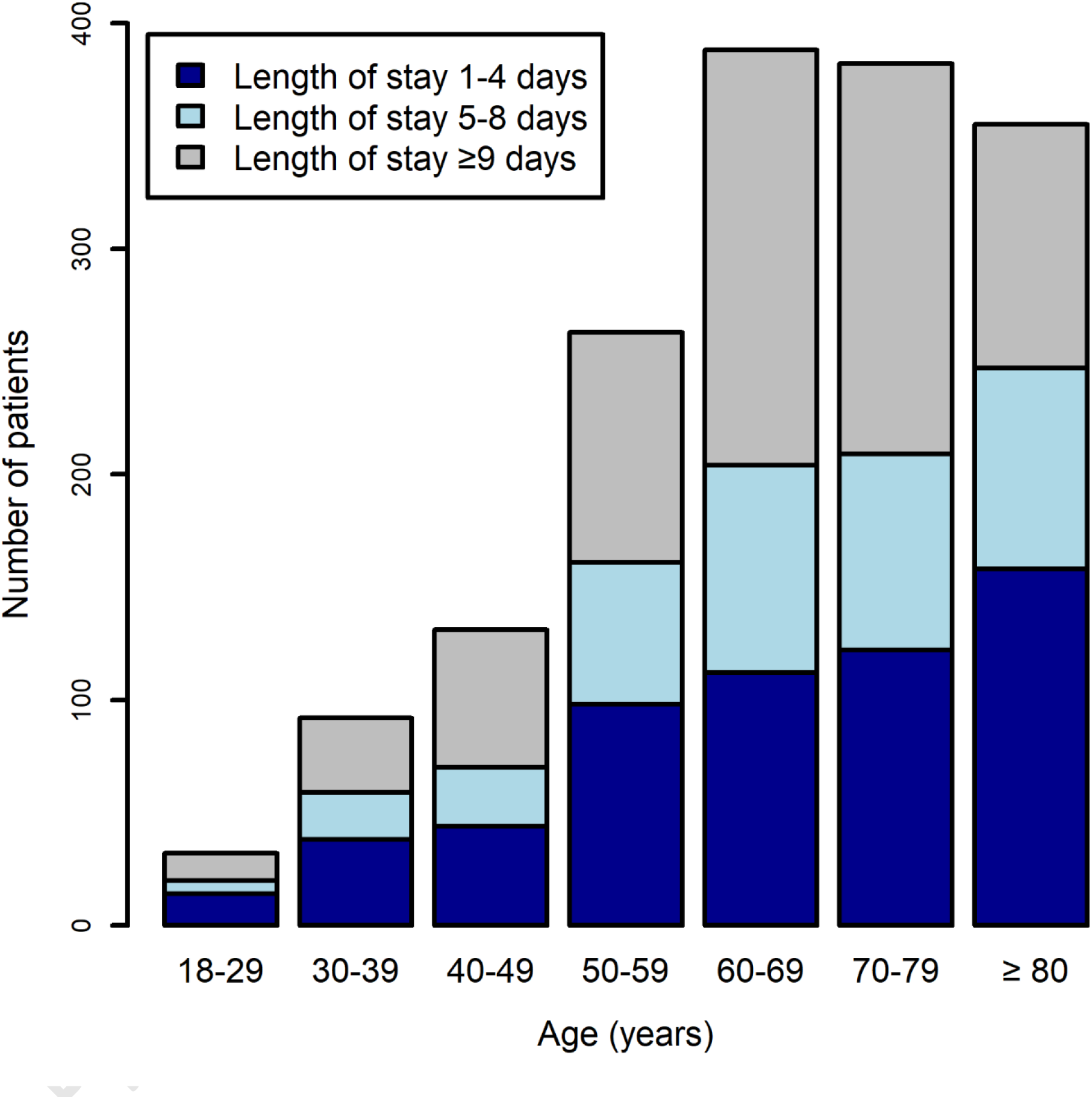
Frequency histogram by age groups for hospital length of stay among severe COVID-19 patients with hospital length of stay of 1-4 days (n=586, 36%), 5-8 days (n=384, 23%), ≥ 9 days (n=673, 41%).

## Discussion

In our cohort the median LOS was markedly shorter than the equivalent endpoint in the ACTT-1 trial (7 days vs. 15 days). This difference raises immediate questions as to whether remdesivir could reduce LOS by the four days used to justify the treatment’s price. We found that only 41% of patients could both receive a 5-day course of remdesivir and have LOS shortened by 4 days or more, while 36% could have their LOS potentially prolonged to complete therapy.

Our evaluation is relevant to the extent that our older-adult multi-morbidity predominant cohort is representative of severe COVID-19; LOS may be even shorter in younger and healthier populations. Our data are from patients hospitalized before dexamethasone became standard of care. Some patients may be prescribed 10 rather than 5 days of remdesivir, which could further prolong LOS (both durations are authorized by the FDA). Whether physicians will keep patients who otherwise could be discharged to complete treatment could not be determined.

Studying shorter remdesivir treatment courses, developing intranasal remdesivir,^6^ and implementing programs to facilitate outpatient intravenous remdesivir administration should be considered.

## Data Availability

Data sharing would be considered under appropriate IRB oversight and approval.

## Author Contributions

Drs Anderson and Baldwin had full access to all of the data in the study and take responsibility for the integrity of the data and the accuracy of the data analysis. **Study concept and design**: Anderson, Bach, Baldwin. **Acquisition, analysis, and interpretation of data**: Anderson, Baldwin. **Drafting of the manuscript**: Anderson, Bach, Baldwin. **Critical revision of the manuscript for important intellectual content:** Anderson, Bach, Baldwin. **Statistical analysis**: Anderson, Baldwin.

